# A Qualitative Assessment of Health Workers’ Knowledge, Perceptions, and Practices on Sudden Unexpected Infant Death in Central Uganda

**DOI:** 10.64898/2026.09.08.26362587

**Authors:** Mary Nyantaro, James Carpenter, Andrew Abaasa, Richard Muhumuza, Sarah Nakamanya, Nabaggala Georgina, Joseph Mugisha, Freddie Mukasa Kibengo, Nambusi Kyegombe

## Abstract

**Introduction:** Health workers (HWs) have a central role in reducing the risk of Sudden Unexpected Infant Death (SUID) through caregiver education and safe-sleep counselling. Yet, awareness and application of evidence-based guidance are inconsistent in primary and maternal–child health settings in low- and middle-income countries (LMICs). This study explored the knowledge, perceptions, and practices of HWs regarding SUID in Uganda.

**Methods:** We conducted a qualitative study with 20 HWs (midwives, nurses, clinicians and medical officers) at three government facilities (two regional referral hospitals and one health centre III) in central Uganda (April–September 2024). Participants were purposively sampled. In-depth interviews were conducted in English or Luganda(a widely spoken local language), audio-recorded, and transcribed into English. Data were analysed thematically using a hybrid inductive–deductive approach supported by NVivo 14.

**Results:** HWs recognized sudden infant deaths but were largely unfamiliar with SUID as a clinical construct. Commonly perceived causes included accidental suffocation, overlay, unsafe sleep positioning and underlying illness, and cultural beliefs. Safe-sleep counselling was infrequent and non-standardized; many HWs recommended prone or side-lying positions to avoid perceived aspiration risk and favoured soft sleep surfaces. Mother infant bed-sharing was considered culturally normative and practical for warmth and monitoring despite acknowledged risks. HWs expressed a need for training and context-adapted guidelines.

**Conclusion:** Participants described gaps and inconsistencies regarding safe sleep and environmental risk factors for SUID. Developing national, context-appropriate safe sleep guidance, integrating pre-service and in-service training, and embedding consistent counselling in maternal–child health services could contribute to risk reduction and consequently prevent infant deaths.

## Introduction

Sudden Unexpected Infant Death (SUID) remains a significant public health concern worldwide (Wilson & Randall, 2021). SUID encompasses all sudden and unexpected deaths of infants, including those whose cause is later explained (e.g., infections, metabolic disorders, or accidental suffocation) and those that remain unexplained after thorough investigation, such as Sudden Infant Death Syndrome (SIDS) (Ferres et al., 2019; Sharma et al., 2024).The triple risk model explains SIDS occurs when an infant with intrinsic vulnerability(genetic, metabolic or exposure to nicotine) is exposed to exogenous trigger such as unsafe sleep environment during a period of critical development. Evidence-based safe sleep practices such as supine sleeping position, the use of firm sleep surfaces, room-sharing without bed-sharing, the avoidance of soft bedding, withholding bed toys and accessories, and avoiding overheating among others have reduced sleep related deaths in developed countries (Diniz et al., 2024; Jullien, 2021; Moon, Carlin, & Hand, 2022). However, in LMICs, including sub-Saharan Africa, SUID burden is likely underrecognized due to limited surveillance, awareness, and lack of national guidance on safe sleep in the routine maternal–child health counselling. (G. Osei-Poku et al., 2023; Osei-Poku et al., 2021; Sun et al., 2025; Winterbach et al., 2021).

Health workers play a critical role in reducing the incidence of SUID through parental education, implementation of safe sleep guidelines, and early identification of risk factors (Doğan & Yılmaz, 2020; Ellis et al., 2022; Landsem & Cheetham, 2022; Ninsiima et al., 2020). However, the extent to which these guidelines are known, perceived, and practiced by frontline health professionals especially in primary care and maternal and child health settings varies considerably across regions and healthcare systems (Vincent et al., 2023). Scooping and systematic reviews on safe sleep have highlighted gaps in health worker knowledge and inconsistent practices regarding infant sleep safety, often influenced by cultural beliefs, lack of training, resource constraints, and conflicting information (Aggelou et al., 2024; Landsem & Cheetham, 2022). These gaps can undermine the effectiveness of public health messaging and may contribute to persistent rates of preventable infant death (Aggelou et al., 2024). Understanding the knowledge base, attitudes, and behaviours of health workers is therefore essential for tailoring interventions, enhancing professional training, and strengthening caregiver counselling strategies.

By contrast, in Uganda, to our knowledge, there is no data on knowledge, perceptions, and practices of health workers regarding SUID prevention. There are no Ugandan safe sleep guidelines. Assessing these dimensions is essential for identifying knowledge gaps, misconceptions, and practices that may hinder the effective dissemination of safe infant care messages. Research on these issues can inform targeted interventions, strengthen health education programmes, and contribute to national strategies aimed at reducing preventable infant deaths in similar resource-constrained settings.

This study is informed by the Social Ecological Model (SEM), which conceptualises health behaviours as being shaped by interactions across multiple levels, including individual, interpersonal, community, organisational, and policy contexts. The SEM was applied retrospectively, once data collection was underway, rather than shaping the initial topic guide, which was built primarily around empirical domains of knowledge, perceived causes and counselling practices. During analysis, the model was used deliberately to organise codes and themes according to the level at which they appeared to operate (individual, interpersonal, community, organisational or policy), which in turn structured the interpretation offered in the Discussion. In relation to SUID, preventive practices such as safe infant sleep are not determined solely by individual knowledge, but are influenced by cultural beliefs, health system structures, professional norms, and the availability of evidence-based guidance. The SEM is therefore particularly relevant for understanding how health workers’ counselling practices emerge within broader social and institutional environments in low- and middle-income settings. This study explored health workers’ knowledge of SUID, perceptions of its causes, and routine infant sleep counselling practices in Uganda, with the aim of identifying barriers and opportunities for integrating safe-sleep guidance into maternal and child health services.

## Methods and materials

### Study design and setting

We conducted a qualitative cross-sectional study (April–September 2024) in central Uganda at Masaka Regional Referral Hospital (Masaka District), Entebbe Regional Referral Hospital (Wakiso District), and Kyamulibwa Health Centre III (Kalungu District). These three facilities serve rural and peri-urban populations. These facilities were selected because they are referral or primary points of care for our existing two infant cohort trials; PRiMe (Nduba et al., 2026) and OPTIMMS (Bijukchhe et al., 2026), and the Kyamulibwa General Population Cohort (GPC) from which SUID deaths was encountered. Entebbe Regional Referral Hospital (ERRH) is a public facility in Uganda’s Central Region funded by the Ministry of Health. It serves a catchment population of about 4 million people across seven districts, including parts of Kampala and airport-related communities. Masaka Regional Referral Hospital (MRRH) is a public facility in south-central Uganda, funded by the Ministry of Health. It is one of Uganda’s designated referral hospitals. MRRH provides specialist, general, preventive, promotive, and rehabilitative services to a catchment population of about 2.6 million people spanning approximately ten districts. Kyamulibwa Health Centre III is a public facility in Kyamulibwa subcounty, Kalungu District, Central Uganda. As a Health Centre III, it provides outpatient care, maternity services, maternal and child health, limited inpatient care, and serves as a referral point for lower-level facilities. It supports an estimated catchment population of about 177,200 people.

**Fig 1:**
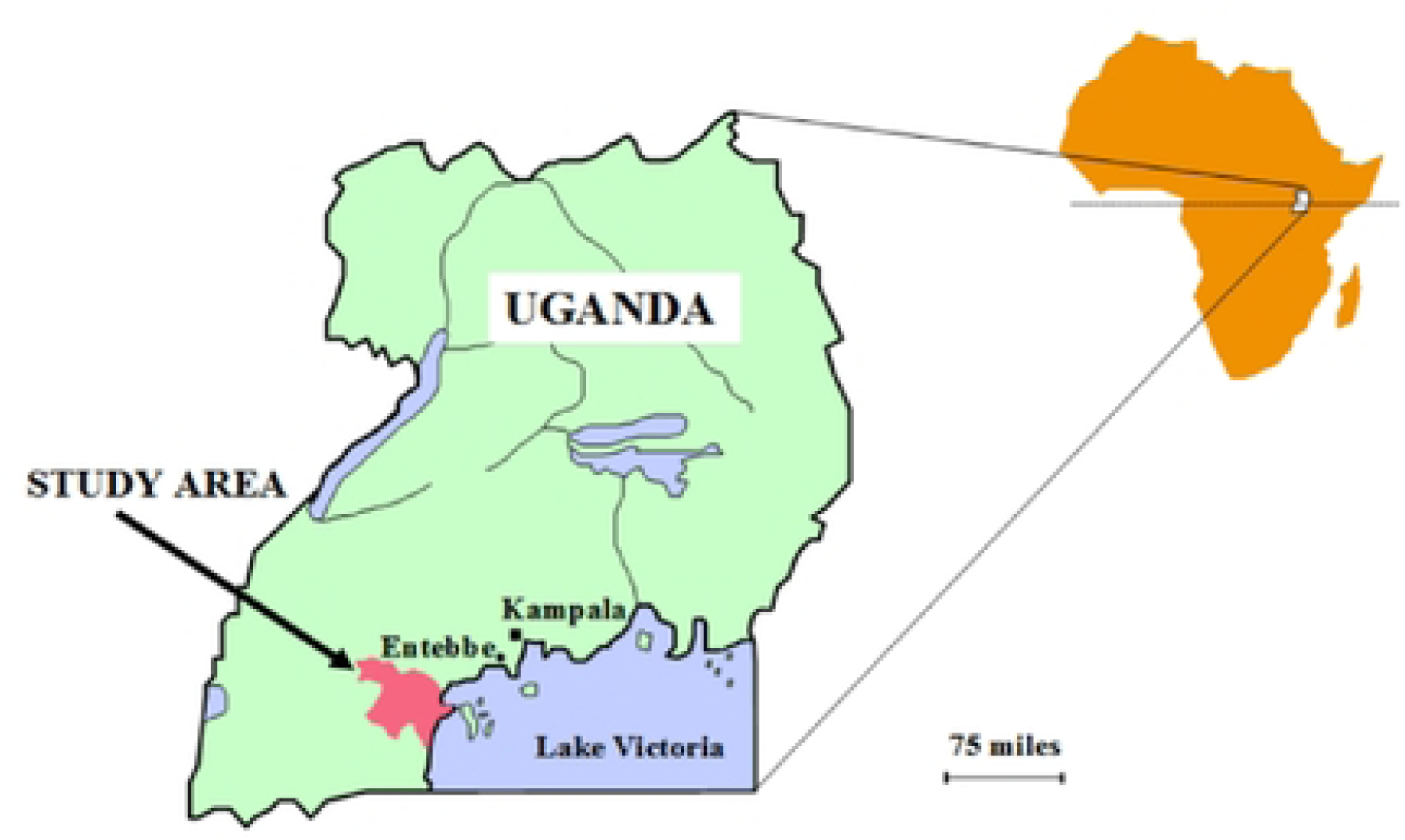
The map of Uganda showing the study sites.

### Participants and sampling

Health workers were purposively sampled to capture variation in cadre (midwives, nurses, clinical officers and medical officers), facility level (regional referral hospital and health centre III) and sex, while also being available and willing to take part during the data collection period. At each facility, the research team approached health workers on duty in maternal and child health departments; the facility in-charge helped to identify staff who were on site and eligible, after which the research team approached potential participants directly to explain the study and seek consent, so as to limit the influence of managers over who ultimately took part. Three health workers(doctor, nurse and midwife) who were approached declined or were unable to participate because of workload. Because recruitment combined purposive and convenience elements, the sample is best understood as reflecting the views of health workers who were both accessible and willing to discuss infant sleep, rather than a representative cross-section of all cadres at these facilities. We recruited 20 HWs (13 female, 7 male) on duty at the time of site visits: Masaka (n=8), Entebbe (n=7), Kyamulibwa (n=5). Eligible cadres included midwives (n=8), nurses (n=5), clinical officers (n=4), and medical officers (n=3) who cared for infants under 12 months of age and provided written informed consent.

### Data collection

We conducted in-depth interviews (IDIs) in a private room at each facility using a semi-structured topic guide in English or Luganda (the main local language), covering domains of knowledge of SUID, perceived causes, counselling content, and sleep practices (positioning, sleep surface, bed-sharing). Of the 20 interviews, fifteen were conducted in English and five in Luganda. Luganda interviews were conducted by bilingual members of the research team (RM,GN and SN) and transcribed directly into English by a bilingual team member; a sample of transcripts was checked against the original audio recordings by a second team member to confirm accuracy, and culturally specific terms or expressions were retained in Luganda within the transcript, with a brief explanatory note, where a direct English equivalent risked losing meaning. Interviews were audio-recorded with consent and lasted 30 to 60 minutes. No repeat interviews were conducted. Participants did not review transcripts. Interviewers documented field notes immediately following interviews. Data collection and analysis proceeded concurrently, and by the seventeenth interview the team judged that no substantially new themes were emerging within the domains covered by the topic guide; the remaining interviews were completed as planned and approved by the ethics committee, and were reviewed to confirm they were consistent with this judgement of thematic sufficiency. Confidentiality was maintained by identifying data with participant ID numbers and storing all information on password protected computers and in locked cabinets at our research office.

### Researcher reflexivity

This study was conducted by a multidisciplinary team based at a medical research institution in Uganda and the United Kingdom. Three team members-two females and one male (SN, GN and RM) were directly involved in data collection. SN, GN and RM are experienced qualitative researchers native to the study area, sharing the local language and familiarity with community norms around infant care. MN is a female clinician with ten years of experience in maternal and child health and did not participate in the data collection because of her position. This combination of clinical and local positioning helped build rapport with health worker participants, who may have viewed the team as credible on clinical matters, but it may equally have encouraged participants to present more guideline concordant or professionally acceptable accounts of their knowledge and practice than they would have shared with a non-clinical or unaffiliated interviewer. At the same time, no member of the data collection team had a prior personal or professional relationship with the specific health workers interviewed, which required active effort to build trust and openness at each site regardless of shared language or background. Five team members-one female and four males (NK, JC, FK, JM and AA) were not involved in data collection or transcription and reviewed coding and interpretation from a more distanced position; their backgrounds in public health, statistics and social science supported a questioning stance towards emerging themes and helped surface assumptions, for example about what counts as a safe sleep practice, that the data collection team may have taken for granted. Reflexive practices, including regular peer debriefing meetings among the research team and discussion of preliminary interpretations at dissemination meetings with colleagues at the research unit, were used throughout to examine how these different positions may have shaped data collection and analysis. This peer debriefing and internal review process was valuable for interrogating the analytical approach and assumptions of the research team, but it does not amount to participant or community validation of the findings; we did not undertake formal member checking with participants, and this is acknowledged as a limitation. Where discussion with colleagues led to a specific change, for example refining the boundary between the sleep surface and bed sharing themes, this is noted in the relevant part of the Results.

### Data management and analysis

Transcripts were imported into NVivo 14. Three members of the research team (SN, RM and MN) independently coded an initial subset of 4 transcripts, chosen to include interviews from each facility and cadre, and met regularly to compare codes and discuss areas of difference. These discussions aimed at a shared, well-reasoned understanding of the data that reflected different analytic perspectives, rather than a single correct interpretation; where the interpretation diverged, this was recorded and revisited against the original transcript. The resulting codebook was then applied by MN to the remaining transcripts, with periodic checks by a second analyst to confirm consistent application. Codes were collated into candidate themes and subthemes. These were reviewed against data extracts and the full corpus, and refined for coherence and distinctiveness. Analysis proceeded concurrently with data collection, allowing the team to probe emergent concepts, such as spiritual explanations for infant death, in later interviews. Coding and theme development were also informed by the Social Ecological Model, used to consider the individual, interpersonal, community, organisational and policy level at which each code or theme appeared to operate. We drew on Guba and Lincoln’s criteria to consider the trustworthiness of the analysis (Ahmed, 2024; Enworo, 2023). Credibility was supported through analyst triangulation, in which the three coders’ independent readings of the initial transcript subset were compared and reconciled as described above. Through peer debriefing, in which the wider research team, including members not involved in data collection, questioned emerging themes at regular meetings. We paid particular attention to accounts that diverged from the majority view, for example the small number of health workers who did offer some form of safe sleep counselling, and considered what these divergent accounts suggested about the conditions under which counselling did or did not occur, rather than setting them aside as outliers. Dependability and confirmability were supported by detailed documentation of the methods. The reflexive stance described above; we have also strengthened the contextual and analytical detail given in the Results. This also included verbatim quotations and description of the settings in which accounts were given, to support the transferability of these findings to comparable primary care and referral settings in Uganda

### Ethical Approvals

Ethical approval: Uganda Virus Research Institute Research Ethics Committee GC/127/997 (Uganda), London School of Hygiene and Tropical Medicine REC 30025 (UK), and Uganda National Council of Science and Technology HS3465ES (Uganda). Participants provided written informed consent for participation and audio recording. Data were de-identified and stored securely.

## Results

### Participant characteristics

Twenty HWs (13 female, 7 male; age 26–59 years) participated across three facilities. Most were midwives and nurses; 75% (n=15) were married and 65% (n=13) had ≥1 child. Professional experience ranged from 2 to 34 years, reflecting a mix of junior and senior healthcare providers and ensuring diverse perspectives on maternal and newborn health service delivery.

**Table 1.** summarizes available characteristics.

| **Characteristic** | **n (%)** |
| --- | --- |
| **Sex: Female** | **13 (65)** |
| **Sex: Male** | **7 (35)** |
| **Age range (years)** | **26–59** |
| **Marital status: Married** | **15 (75)** |
| **Has $\geq 1$ child** | **13 (65)** |
|  | **8 (40)** |
| **Facility: Entebbe RRH** | **7 (35)** |
| **Facility: Kyamulibwa HC III** | **5 (25)** |

### Emergent Themes

We identified three interlinked domains, developed from the data through the coding process described above rather than mapped directly onto the topic guide, although the topic guide’s focus on knowledge, perceived causes and counselling naturally shaped the broad areas explored: (i) knowledge of SUID as a clinical construct, set against lived awareness of sudden infant deaths in the community; (ii) perceived causes of SUID, spanning suffocation and overlay, sleep posture, cultural and spiritual explanations, unnoticed illness, and the sleep surface and environment; and (iii) practices and counselling, covering how, when and to whom health workers offered safe sleep advice, and how this advice was shaped by beliefs about sleep positioning, bed sharing and sleep surfaces. We observed substantial consistency in themes across cadres, facility types and study sites.

## 1. Knowledge of SUID

A small number of health workers, generally the more senior clinical officers and medical officers who had trained more recently or had exposure to paediatric rotations, were familiar with the term SUID or similar guideline language and could describe it in general terms as an unexplained infant death during sleep; however, even these participants had limited knowledge of specific risk factors or recommended prevention practices, and none had received dedicated training on the topic. Most participants became aware of sudden infant deaths through community experience rather than formal clinical training

> *“I have never heard of that condition as a medical condition. I have heard of cases of babies dying in their sleep in the community though.” Nurse 01 female Masaka*

HWs expressed uncertainty about aetiology and a strong desire for updated training and practical guidance.

> *“As health workers we get new information every day and we need to talk about the major causes of death in infants. However, we need information on SUID and a poster. We need training as health workers.” Midwife 04 female, Masaka*

## 2. Perceived Causes of SUID

Across the sample, perceptions of suffocation, sleep posture, cultural attributions, unnoticed illness and sleep surface risk did not differ in any systematic way by cadre, facility or years of experience; the same set of concerns and explanations recurred among midwives, nurses, clinical officers and medical officers at all three sites. We report this explicitly here because it shapes how the following sub-themes are presented: rather than organising the account by professional group, we present the perceived causes as a shared repertoire drawn on across cadres, while noting individual variation and any exceptions within the extracts themselves.

### 2.1. Sleep related risks factors

#### 2.1.1. Suffocation

HWs frequently emphasized accidental suffocation especially during bed-sharing when a fatigued or intoxicated caregiver might roll onto the infant; other scenarios included bottle-feeding in bed and loose bedding obstructing the airway:

> *“A drunk mother ….can forget that she is sleeping with an infant…. infant might turn and lay in a bad position and suffocates.” Clinician 05 male Masaka*
>
> *“The infant’s face may get stuck in clothes and suffocates. Also, the drinks might choke him/her…..mothers, after giving the infant the feeding bottle…..The infant might fail to swallow the drink or s/he falls asleep and chokes.” Midwife 01 female, Kyamulibwa*

#### 2.1.2. Sleep posture

Most HWs viewed prone and side-lying positions as safer for breathing and for preventing aspiration, expressing concerns about supine sleep. The accounts highlighted the need for caregivers to be vigilant in monitoring infants’ breathing and positioning, ensuring that the sleep environment does not allow clothes, bedding, or the surface itself to block the airway.

> *“The side sleeping is important if an infant can change. Sleeping while facing down can cause suffocation. Always put a towel and pillow to maintain the head on the side.” Nurse 02 female, Entebbe*

#### 2.1.3. Sleep surfaces and environment

Participants considered soft mattresses, polythene covers, and loose clothing as hazards that could block the airway. some recommended moderately soft surfaces for infant comfort.

> *“Facing downward is when sometimes the polythene also gets the opportunity to suffocate the infant as the infant struggles to get off. It is better for the infant to sleep face side not down.” Nurse 02 female, Kyamulibwa*

### 2.2. Cultural beliefs

Cultural interpretations were salient, including spiritual or moral attributions (e.g., extramarital relations, witchcraft) as causes of sudden deaths. They noted some community members believed an infant may die if a parent engages in extramarital sexual relations particularly if the father returns home and touches the infant, or if a breastfeeding mother has intercourse with another man. Some of the health workers expressed doubt about the scientific validity of these claims.

> *“I do not know about how true that is, but people say that if an infant’s father commits adultery and comes back home and touches the infant, he can cause this condition, but I do not know the science behind it. What I know is that some deaths are abrupt and people choose to attach that as a reason.” Midwife 06 female, Kyamulibwa*
>
> *“Actually, that’s when the infant is still breastfeeding. Then the mother has intercourse with another man. There is a high chance of the infant dying. But I don’t think that’s true.” Medical officer 02 male, Entebbe*
>
> *Putting an infant in a separate room. Evil spirits could come and take your infant. I would advise the mother to keep the infant in the cradle for those who sleep badly. Like the infant of King Solomon. Medical Officer 2 male, Entebbe*

### 2.3. Unnoticed illness

HWs cited undiagnosed congenital heart disease, genetic disorders, or early respiratory illness as possible contributors that may not present with clear warning signs. They noted that caregivers often lack the knowledge or clinical awareness to recognise subtle symptoms, leading them to assume the infant is healthy until an abrupt and unexplained death occurs. In these situations, the absence of visible warning signs were described to leave families confused and searching for answers, while the underlying cause may have been a hidden medical condition that quietly progressed without detection. This pattern, of health education being directed mainly towards commonly encountered, immediately life-threatening conditions rather than towards SUID or safe.

> *“Congenital abnormalities especially involving the heart maybe unnoticed for some time and the only sign is infant sudden death. The infant just dies, and you keep questioning yourself what could have killed this infant.” Midwife 06 female, Entebbe)*

## 3. Practices and counselling

### 3.1. Health Education scope

Health education provided by health workers mainly addressed infant health problems that are commonly encountered and seen as immediately life-threatening, including pneumonia, malaria, anaemia, and nutrition. In contrast, no attention was given to sudden unexpected infant death (SUID) or safe sleep practices. This focus appears to be influenced by the realities of busy and resource-limited clinical settings, where health workers prioritize conditions they routinely manage and feel most equipped to address. As one clinician explained, health education was not consistently offered to all mothers but was instead directed toward those considered to be at greatest immediate risk of contracting those common illnesses. This view was held by all the participants from rural and urban settings.

> *“The nurses are the ones that usually educate mothers. We usually tell them some of the topics. I do not want to lie, the health education is not done to most mothers. I educate a few. As a health worker, I usually educate to save life. I educate those of severe pneumonia, anaemia and lack of proper care”. Clinician 05 male, Masaka*
>
> *“SIDS is never discussed. I have not heard about anyone telling mothers how to safely put an infant to sleep”. Clinician 04 male, Kyamulibwa*

We further examined the extent to which sleep practices were addressed during health education talks provided to parents at antenatal and other facility visits. Health workers reported that guidance on infant sleep practices was rarely included in routine education. However, when prompted, they shared their perspectives on specific aspects of infant sleep, including sleeping position, sleep surface, and co-sleeping.

> *“We don’t emphasize on the sleeping practices of infants as an important thing. Most time we talk about other things like breast feeding. But we encourage the mothers to sleep with their infants.” Nurse 02 female, Masaka*

### 3.2. Sleeping position

Most HWs advised prone or side-lying sleep, occasionally with supports (e.g., rolled towels) to maintain position and avoid perceived choking. Supine sleep was sometimes advised immediately post-delivery for cord care airflow, then replaced by prone/side-lying afterward.

> *“We usually prefer the infant to lie on the stomach. It helps the infant to sleep well and get enough rest, and even breathing becomes okay. If you try to make an infant lie on the back, he/she might get choked with saliva or anything.” Midwife 06 Female, Entebbe*
>
> *“More especially when the infant has just been delivered, people have a practice of making the infant sleep facing down but we tell them when the umbilical is still wet, according to what we studied, an infant should face up to have enough air to dry the umbilical cord… According to hearsay, it’s because when the infant sleeps facing down, s/he can sleep for some good time compared to the one that is facing up”. Nurse 06 male, Masaka*

### 3.3. Sleep surface

Many recommended soft/flat mattresses to improve comfort; fewer emphasized firm, uncluttered surfaces. Some mentioned polythene coverings as a risk. When participants were probed further, what they described as a soft surface, this was most often a foam mattress. This would be sometimes have additional padding or a blanket.

> *“The surface should be flat and soft. The soft one is okay, not the hard one. Because you can’t put an infant on something hard, like a table. So, it should be a flat and soft mattress.” Nurse 01 female, Entebbe*
>
> *“The infant sleep surface should be soft. The infant’s body is small and soft (fragile). The hard surface may make the infant disfigured. Keep changing his/her position”. Nurse 02 female, Masaka*

### 3.4. Co sleeping/ Bed sharing

Bed sharing was described as a practice influenced by both cultural expectations and practical realities. Participants noted that limited household space, lack of separate infant sleeping bed, the convenience of nighttime breastfeeding, the need to keep infants warm, and the ability to monitor infants more closely all contributed to bed sharing. These findings suggest that bed sharing is often a response to socioeconomic and caregiving constraints.

> *“I would advise these mothers who sleep with their infants to be careful. Because some people sleep and turn and sleep on the infant. So, if these infants are sleeping, parents have to be very cautious. Some infants will turn and sleep on their faces. And the next thing, these infants will not be breathing…” Medical doctor 02 male, Entebbe*

Furthermore, some of the health workers were against having an infant sleep in their separate room as this complicates checking on the infant. A few had spiritual beliefs that some sort of evil spirits would come and take the infant. One went ahead to cite the Bible where a infant died during sleep

> *“Putting the infant in a separate room, evil spirits could come and take your infant. I would advise the mother to keep the infant in the cradle for those who sleep badly. Like the infant of King Solomon.” Nurse 2 female, Masaka*

## Discussion

This study highlights three key findings. First, the absence of formal training, national guidance, and routine inclusion of SUID in maternal and child health education meant that healthcare workers relied largely on experiential learning, fragmented biomedical knowledge, and local explanations, resulting in inconsistent understanding of SUID (Moon et al., 2016; Parks et al., 2017; Shapiro-Mendoza et al., 2023). Second, these knowledge gaps contributed to well-intentioned but potentially unsafe recommendations. The persistence of prone and side-sleep advice suggests that concerns about aspiration continue to outweigh adherence to contemporary safe-sleep evidence. Similar misconceptions have been reported across both high- and low-income settings, indicating that training alone may be insufficient without addressing the cognitive models through which health workers understand infant airway protection. Third, infant sleep practices were strongly shaped by socioeconomic and caregiving realities. Bed sharing and use of softer sleep surfaces reflected housing constraints, poverty, thermal comfort needs, breastfeeding, and family sleeping arrangements, underscoring the need for locally adapted safe-sleep guidance that translates evidence-based recommendations into practical options for Ugandan households(Carpenter et al., 2013; Moon, Carlin, Hand, et al., 2022).

Viewed through the Social Ecological Model (SEM), these findings demonstrate influences operating across multiple levels. At the individual level, healthcare workers reported limited knowledge of SUID and misconceptions about infant sleep positioning. Interpersonal influences included family caregiving practices and beliefs regarding infant monitoring. Community-level factors encompassed cultural and spiritual explanations for infant death, while organisational and policy-level influences were reflected in the absence of training, guidelines, and standardised counselling protocols.

These findings are consistent with studies from both low- and high-income settings showing gaps in provider knowledge and variation in counselling practices, often shaped by training exposure, resource limitations, and cultural norms (G. K. Osei-Poku et al., 2023; Shapiro-Mendoza et al., 2023). Counselling priorities appeared influenced by the realities of busy, resource-constrained clinical settings. At the same time, reported practices diverged from global recommendations advocating supine sleep on a firm, flat, uncluttered surface, room sharing without bed sharing, and avoidance of soft bedding and overheating. The persistence of bed sharing highlights the need for context-sensitive, harm-reduction approaches rather than strict prohibitions.

Most healthcare workers understood sudden infant deaths as unexpected deaths occurring during sleep in otherwise healthy infants. However, knowledge was largely informal and community-derived rather than acquired through structured training, mirroring findings from other low- and middle-income settings (Ndu, 2016). The limited integration of SUID into maternal and child health education represents a missed prevention opportunity, given the central role of healthcare providers in promoting safe-sleep practices (Moon et al., 2016; Ndu, 2016).

At the organisational level, the absence of national guidelines and formal training strongly influenced practice. Consequently, healthcare workers often relied on personal experience and culturally embedded beliefs, contributing to inconsistent and sometimes unsafe counselling. This reflects SEM principles, whereby organisational and policy environments shape frontline practice.

Participants identified suffocation, cultural factors, and underlying medical conditions as causes of SUID. Suffocation was commonly linked to unsafe sleep environments, bed sharing, caregiver fatigue, and inappropriate bedding, consistent with evidence identifying accidental suffocation and overlay as important contributors to sleep-related infant deaths (Hauck & Blackstone, 2022; Hauck et al., 2008; Moon et al., 2016). Some participants also attributed deaths to undiagnosed medical conditions, reflecting biomedical understandings that a proportion of cases may be associated with underlying disease processes (Sharma et al., 2025). Cultural explanations, including witchcraft and moral transgressions, were also reported, consistent with findings from other African settings (G. K. Osei-Poku et al., 2023).

Evidence suggests that risks associated with bed sharing are context dependent and increase substantially in the presence of hazards such as parental smoking, alcohol or drug use, prematurity, low birth weight, soft bedding, and sofa or armchair sleeping (Carpenter et al., 2013; Moon et al., 2016). Consequently, safe-sleep messaging in Uganda may benefit from context-sensitive harm-reduction approaches that clearly identify high-risk circumstances rather than treating all bed sharing as equally hazardous.

At the policy level, national safe-sleep guidance should be integrated within existing maternal and newborn health programmes and delivered through established platforms such as antenatal care, postnatal services, and community health worker outreach. Such guidance must take account of the housing conditions, caregiving arrangements, and resource constraints identified in this study. While these findings help identify priorities for policy development and local adaptation, they cannot determine the national feasibility, acceptability, or effectiveness of specific recommendations, which will require further research and stakeholder engagement, including with the Ministry of Health.

This study has several limitations. Selection bias may have occurred because some facilities were linked to ongoing cohort studies and participation depended on staff availability. The small sample limited comparisons across cadres and facilities, and recruitment through facility leadership may have influenced participant selection. Findings were based on self-reported knowledge and practices, which may not reflect actual counselling behaviour, and translation from Luganda to English may have resulted in some loss of nuance. Caregiver and policymaker perspectives were not included. As a qualitative study, the findings are intended to be transferable to similar contexts rather than statistically generalisable.

## Conclusion

Healthcare workers recognise sudden infant deaths but lack consistent knowledge and counselling approaches. These findings identify clear priorities for co-developing national safe sleep guidance, in-service and pre-service training, and standardised counselling approaches with health workers, policymakers and communities in Uganda, and for evaluating whether such approaches are acceptable, feasible and, ultimately, may strengthen the consistency of safe-sleep messaging and represents a plausible strategy for reducing preventable sleep-related infant mortality.

## Data Availability

Anonymised, excerpted data will be held in LSHTM’s data repository https://datacompass. lshtm.ac.uk/ and can be requested through this site by legitimate researchers. Requests will be considered on a case-by-case basis with criteria for access including possession of ethics approval, data use agreement, purpose etc. Response times are usually 2-3 weeks. All data requests for select excerpts to clarify any ambiguities should be sent to the First author Mary Nyantaro (Mary.nyantaro1@lshtm. ac.uk), or contact research data management at.

## Authors’ contributions

- Conceptualization: MN, JC, AA, FK, JM, NK
- Methodology: MN, RM, SN, GN, AA, JC, NK
- Investigation (data collection): MN, GN, RM, SN
- Formal analysis: MN, RM, SN
- Writing – original draft: MN
- Writing – review & editing: RM, SN, GN, FK, JM, AA, JC, NK
- Supervision: AA, JC, NK
- Funding acquisition: MN, MRC/UVRI & LSHTM Uganda Research Unit, Royal Society of Tropical Medicine and Hygiene

## Acknowledgements

We acknowledge the contributions of MRC/UVRI and LSHTM Uganda Research Unit and Royal Society of Tropical Medicine and Hygiene Early Career Research Grant for the support. We are particularly grateful to all the study participants without forgetting the mobilisers for the time and information they shared with us.

## Availability of data and materials

Data will not be shared publicly due to the data-sharing policy of the LSHTM requiring a prior data-sharing agreement. However, the dataset containing the data supporting the study findings in this report can be obtained upon request from the corresponding author.

## Consent for publication

Not applicable

## Reference

Aggelou, M., Metallinou, D., Dagla, M., Vivilaki, V., & Sarantaki, A. (2024). Evaluating Educational Patterns and Methods in Infant Sleep Care: Trends, Effectiveness, and Impact in Home Settings—A Systematic Review. Children, 11. 10.3390/children11111337

Ahmed, S. K. (2024). The Pillars of Trustworthiness in Qualitative Research. *Journal of Medicine*, Surgery, and Public Health. 10.1016/j.glmedi.2024.100051

Bijukchhe, S. M., Marchevsky, N. G., Kibengo, F., Sharma, A. K., Basi, R., Cantrell, L., Chuke, S., Curtis, B., Diavatopoulos, D. A., Eordogh, A., Gautam, M. C., Gurung, M., den Hartog, G., Kattel, H. P., Kc, S., Kelly, D., Kelly, S., Li, G., Maskey, P.,…Pollard, A. J. (2026). Optimising DTwP-containing vaccine infant immunisation schedules in Uganda and Nepal (OptImms): two open-label, non-inferiority, randomised controlled trials. Lancet Infect Dis, 26(7), 726–738. 10.1016/s1473-3099(26)00053-8

Carpenter, R., McGarvey, C., Mitchell, E. A., Tappin, D. M., Vennemann, M. M., Smuk, M., & Carpenter, J. R. (2013). Bed sharing when parents do not smoke: is there a risk of SIDS? An individual level analysis of five major case–control studies. BMJ open, 3(5), e002299.

Diniz, J. D., Soares, B. S., Figueiredo, A. V., Silva, G. P. S., Santos, L. P., & Soares, A. R. (2024). Relationship between the position adopted by the baby during sleep and the prevention of sudden infant death. IV Seven International Congress of Health. 10.56238/homeivsevenhealth-046

Doğan, P., & Yılmaz, H. B. (2020). Nurse’s Role in Reducing the Risk of Sudden Infant Death Syndrome and Creating a Safe Sleep Environment. 7, 75–79. 10.4274/jtsm.galenos.2020.07379

Ellis, C., Pease, A., Garstang, J., Watson, D., Blair, P., & Fleming, P. (2022). Interventions to Improve Safer Sleep Practices in Families With Children Considered to Be at Increased Risk for Sudden Unexpected Death in Infancy: A Systematic Review. Frontiers in Pediatrics, 9. 10.3389/fped.2021.778186

Enworo, O. (2023). Application of Guba and Lincoln’s parallel criteria to assess trustworthiness of qualitative research on indigenous social protection systems. Qualitative Research Journal. 10.1108/qrj-08-2022-0116

Ferres, J. L., Anderson, T., Johnston, R., Ramirez, J., & Mitchell, E. (2019). Distinct Populations of Sudden Unexpected Infant Death Based on Age. Pediatrics, 145. 10.1542/peds.2019-1637

Hauck, F. R., & Blackstone, S. R. (2022). Maternal Smoking, Alcohol and Recreational Drug Use and the Risk of SIDS Among a US Urban Black Population. Front Pediatr, 10, 809966. 10.3389/fped.2022.809966

Hauck, F. R., Signore, C., Fein, S. B., & Raju, T. N. (2008). Infant sleeping arrangements and practices during the first year of life. Pediatrics, 122 *Suppl 2*, S113–120. 10.1542/peds.2008-1315o

Jullien, S. (2021). Sudden infant death syndrome prevention. BMC pediatrics, 21. 10.1186/s12887-021-02536-z

Landsem, I., & Cheetham, N. (2022). Infant sleep as a topic in healthcare guidance of parents, prenatally and the first 6 months after birth: a scoping review. BMC Health Services Research, 22. 10.1186/s12913-022-08484-3

Moon, R., Carlin, R., & Hand, I. (2022). Evidence Base for 2022 Updated Recommendations for a Safe Infant Sleeping Environment to Reduce the Risk of Sleep-Related Infant Deaths. Pediatrics, 150 1. 10.1542/peds.2022-057991

Moon, R., Carlin, R., Hand, I., Jawdeh, E. A., Colvin, J., Goodstein, M., Hauck, F., Hwang, S., Cummings, J., Aucott, S., Guillory, C., Hudak, M., Kaufman, D., Martin, C., Pramanik, A., Puopolo, K., Bundock, E., Kaplan, L., Brown, S.,…Couto, J. (2022). Sleep-Related Infant Deaths: Updated 2022 Recommendations for Reducing Infant Deaths in the Sleep Environment. Pediatrics. 10.1542/peds.2022-057990

Moon, R. Y., Darnall, R. A., Feldman-Winter, L., Goodstein, M. H., Hauck, F. R., & Syndrome, T. F. o. S. I. D. (2016). SIDS and other sleep-related infant deaths: evidence base for 2016 updated recommendations for a safe infant sleeping environment. Pediatrics, 138(5).

Ndu, I. K. (2016). Sudden infant death syndrome: an unrecognized killer in developing countries. Pediatric Health Med Ther, 7, 1–4. 10.2147/phmt.S99685

Nduba, V., Mathebula, M., Nankabirwa, V., Dempster, M., Mhimbira, F., Sabi, I., Wajja, A., Cotton, M. F., Barnabas, S., Adegnika, A. A., Kaufmann, S. H. E., Brückner, S., May, M., Höß, C. W., Revathy, M., Niranjan, V., Bhargava, A., Gupta, M., Kapse, D.,…Niranjan, V. (2026). Comparison of VPM1002 with BCG in the prevention of tuberculosis in newborn infants: a multicentre, double-blind, randomised, phase 3, non-inferiority trial. The Lancet Infectious Diseases. 10.1016/S1473-3099(26)00374-9

Ninsiima, A., Coene, G., Michielsen, K., Najjuka, S., Kemigisha, E., Ruzaaza, G., Nyakato, V., & Leye, E. (2020). Institutional and contextual obstacles to sexuality education policy implementation in Uganda. Sex Education, 20, 17–32. 10.1080/14681811.2019.1609437

Osei-Poku, G., Mwananyanda, L., Elliott, P., MacLeod, W., Somwe, S., Pieciak, R., & Gill, C. (2023). The apparent burden of unexplained sudden infant deaths in Lusaka, Zambia: Findings from analysis of verbal autopsies. Gates Open Research. 10.12688/gatesopenres.14303.1

Osei-Poku, G., Thomas, S., Mwananyanda, L., Lapidot, R., Elliott, P., MacLeod, W., Somwe, S., & Gill, C. (2021). A systematic review of the burden and risk factors of sudden infant death syndrome (SIDS) in Africa. Journal of Global Health, 11. 10.7189/jogh.11.04075

Osei-Poku, G. K., Mwananyanda, L., Elliott, P. A., MacLeod, W. B., Somwe, S. W., Pieciak, R. C., Hamapa, A., & Gill, C. J. (2023). Qualitative assessment of infant sleep practices and other risk factors of sudden infant death syndrome (SIDS) among mothers in Lusaka, Zambia. BMC pediatrics, 23(1), 245.

Parks, S. E., Erck Lambert, A. B., & Shapiro-Mendoza, C. K. (2017). Racial and Ethnic Trends in Sudden Unexpected Infant Deaths: United States, 1995-2013. Pediatrics, 139(6). 10.1542/peds.2016-3844

Shapiro-Mendoza, C. K., Woodworth, K. R., Cottengim, C. R., Erck Lambert, A. B., Harvey, E. M., Monsour, M., Parks, S. E., & Barfield, W. D. (2023). Sudden Unexpected Infant Deaths: 2015-2020. Pediatrics, 151(4). 10.1542/peds.2022-058820

Sharma, S., Whitney, R., Chowdhury, S., & Ramachandrannair, R. (2024). Sudden unexpected infant death, sudden unexplained death in childhood, and sudden unexpected death in epilepsy. Developmental Medicine and Child Neurology, 67, 734–739. 10.1111/dmcn.16226

Sharma, S., Whitney, R., Chowdhury, S. R., & Ramachandrannair, R. (2025). Sudden unexpected infant death, sudden unexplained death in childhood, and sudden unexpected death in epilepsy. Developmental Medicine & Child Neurology, 67(6), 734–739.

Sun, Y., Peng, H., Chen, Q., Qin, L., Ren, Y., & Cheng, Y. (2025). Global, regional, and national burden of sudden infant death syndrome, 1990–2021: a comprehensive analysis of GBD 2021 data with insights into the impact during the COVID-19 pandemic. Frontiers in Pediatrics, 13. 10.3389/fped.2025.1606910

Vincent, A., Chu, N. T., Shah, A., Avanthika, C., Jhaveri, S., Singh, K., Limaye, O., & Boddu, H. (2023). Sudden Infant Death Syndrome: Risk Factors and Newer Risk Reduction Strategies. Cureus, 15. 10.7759/cureus.40572

Wilson, A., & Randall, B. (2021). Trends in Rates of Sudden Unexpected Infant Death (SUID): Hopes for Prevention. South Dakota medicine : the journal of the South Dakota State Medical Association, 74 *5*, 220–226. https://consensus.app/papers/trends-in-rates-of-sudden-unexpected-infant-death-suid-wilson-randall/d47f3d437f4f567b8d26ee3e031c1473/

Winterbach, M., Hattingh, C., & Heathfield, L. (2021). Retrospective study of sudden unexpected death of infants in the Garden Route and Central Karoo districts of South Africa: Causes of death and epidemiological factors. South African Journal of Child Health. 10.7196/sajch.2021.v15i2.01729

